# Nicotine metabolite ratio as an informed biomarker to optimize acupuncture for smoking cessation: randomised controlled trial

**DOI:** 10.1101/2024.10.23.24315983

**Authors:** Shu-min Chen, Jia Ji, Chai xin, Yang Li, Wuyuan, Jin Chang, MiaoZhang, Zhen-yu Liu, Chao-ren Tan, Jin-sheng Yang, Zhao Liu, Ying-ying Wang

## Abstract

**Objective:** To evaluate the effectiveness of using nicotine metabolite ratio (NMR) as an informed biomarker to optimize acupuncture for smoking cessation.

**Methods:** This was a prospective, two-arm, open-label randomized controlled trial. Participants were recruited and classified as slow or normal metabolizers based on their NMR values, and were randomly assigned to high-frequency or low-frequency acupuncture groups. Acupuncture sessions were held 3 to 5 times per week for 8 weeks. Minimum behavioral support was provided. The primary outcome was the 12-week continuous abstinence rate assessed by urine cotinine levels. Secondary outcomes included point abstinence, treatment adherence, and adverse reactions.

**Results:** Between September 2018 and April 2022, 220 participants were included. Among them, 211 (95.9%) were male, and the mean (SD) age was 48.5 (11.6) years. With 0.31 as the NMR cut-off value, 70 (31.8%) were divided as normal metabolizers and 150 (68.2%) as slow metabolizers. The validated 3-month sustained abstinence was 17.3% (19/110) in the high-frequency acupuncture group, which was significantly higher than 8.2% (9/110) in the low-frequency acupuncture group (OR=3.89, 95% CI: 1.36, 8.11). In slow metabolizers, the validated 3-month sustained abstinence rate was 19.0% (15/79) in the high-frequency acupuncture, which was higher than 7.0% (5/71) in low-frequency acupuncture (OR=3.53, 95% CI: 1.03, 6.13). In normal metabolizers, the validated 3-month sustained abstinence rates were 12.9% (4/31) in the high-frequency acupuncture group and 10.3% (4/39) in the low-frequency acupuncture group, with high-frequency acupuncture non-inferior to low-frequency acupuncture (OR=1.11, 95% CI: 0.24, 5.17). Treatment adherence was high in both groups, and adverse reactions were infrequent. No serious adverse events were recorded.

**Conclusion:** This pioneering study demonstrated that NMR might be used as a biomarker to optimize acupuncture treatments for smoking cessation, particularly among slow metabolizers. This study provides valuable insights into personalized smoking cessation strategies and highlights the potential of integrating acupuncture with other interventions for improved outcomes.

**Trial Registration:** Chinese Clinical Trials Registry (No.ChiCTR1800018196), Registered 4 September 2018, (chictr.org.cn).

## Introduction

Despite substantial reductions in tobacco use since the 1960s, the prevalence of tobacco use has remained high in the China for the past decade [1]. Tobacco use brings huge health and economic losses to Chinese people. A recent large-scale population study showed that smokers had significantly higher risks of 56 diseases and 22 causes of death [2]. As a result, nearly 20% of Chinese adult men and about 3% of women will die of smoking, unless effective cessation strategies are implemented widely [3].

Acupuncture, as a complementary and alternative therapy, has been utilized for smoking cessation for over 40 years internationally [4]. Our previous meta-analysis indicated that acupuncture is an effective treatment for smoking cessation, with significant improvement in both short-term (4 weeks, RR 2.37, 95% CI 1.41 to 3.97) and long-term cessation rates (beyond 6 months, RR 2.66, 95% CI 1.50 to 4.70) [5]. Particularly, our previous studies have suggested that acupuncture’s abstinence rates are comparable to those of nicotine replacement therapy and more cost-effective than conventional first-line medications [6–7]. These findings support the potential of acupuncture as an affordable and effective smoking cessation tool.

However, enhancing the efficacy of acupuncture for smoking cessation remains a significant challenge. Recently, nicotine metabolite ratio (NMR) has emerged as a promising biomarker to optimize cessation treatments [8]. Previous studies showed that smokers with high NMR levels (normal metabolizers) tend to have faster nicotine metabolism, leading to more frequent nicotine supplementation and lower success rates in quitting smoking [9–10]. Another randomized controlled trial (RCT) showed that varenicline was more effective than nicotine replacement monotherapy for normal metabolizers but not slow metabolizers [11]. In addition, NMR was also used as a predictor to adjust the dose of smoking cessation medication [12].

Despite the potential utility of NMR, no studies to date have explored its feasibility and possibility of NMR as a biomarker to optimize acupuncture for smoking cessation. To address the evidence gap, we conducted the first randomized controlled trial (RCT) to evaluate the association between acupuncture for smoking cessation and NMR. This pioneering study not only filled a significant gap in the literature but also set the groundwork for targeted and effective acupuncture-based cessation strategies.

## Methods

### Design

This was a prospective 2-arm, open-label, randomized controlled trial. The trial was approved by the Chinese Clinical Trial Ethics Review Committee (Grant No.ChiECRCT-20180135) and registered in the Chinese Clinical Trials Registry (Reg. No. ChiCTR1800018196). All participants provided signed informed consent before participation in the study.

### Participants

This study was conducted at Institute of Acupuncture and Moxibustion, China Academy of Chinese Medical Sciences (Beijing, China), and Wangjing Hospital of China Academy of Chinese Medical Sciences (Beijing, China). Participants were recruited via trial sites, local newspapers, community events, websites, and referrals from other medical institutions.

Participants were included if they were age 18 to 70 years; smoked at least 10 cigarettes per day for at least 1 years, had expired air carbon monoxide (CO) > 6 parts per million (ppm), had urine cotinine concentrations <100 ng/mL, and were motivated to stop smoking. Exclusion criteria included pregnancy or breastfeeding, use of stop-smoking medication during the previous 30 days, history of severe psychiatric illness, unwillingness to use acupuncture for smoking cessation, and current diagnosis of cancer or in remission from cancer for less than 1 year.

### Randomization and blinding

Randomization was conducted via a central randomization system for clinical research (http://210.73.61.115), which was used in our previous study [6]. Randomization sequences were generated using Proc Plan in SAS, version 9.3 (SAS Institute), with trial sites as the stratification factor and a block length of 5. After logging into the website, staff entered participants’ sex, age, and Fagerstrom Test for Cigarette Dependence (FTCD) score, and the system generated each participant’s identification number and treatment allocation via stratified block randomization. The study statistician was masked to treatment codes until the analysis of primary outcome was completed.

### Procedures

Potential participants contacted the local study sites to obtain study details and for eligibility checks. Eligible participants were invited for a baseline visit. At the visit, after they provided a CO reading via a Bedfont Micro Smokerlyzer, and urine cotinine reading by NicAlert strips, their eligibility was confirmed, study details were discussed, and participants signed the informed consent form. They then filled in study questionnaires. After that, participants set up their target quit date (TQD), normally 2 weeks after the baseline visit.

Participants were then randomized into 1 of the two groups. They were also instructed to join a WeChat group for motivational support. Participants were then seen at the study center monthly for 6 months. At each visit, study forms were completed and CO readings were taken. Each follow-up visit took approximately 10 minutes. During the study period, the acupuncture was provided free of charge. The study started recruitment in September 2018 and completed all follow ups in April 2022.

### Interventions Acupuncture

The acupuncture applied in this study was traditional Chinese acupuncture. Selection of Acupoints: Baihui(GV 20), Lieque(LU 7), Hegu(LI 4), Zusanli(ST 36), Sanyinjiao(SP 6), Taichong(LR 3); add Yintangb(EX-HN 3) when there are withdrawal symptoms such as cough and runny nose and dry eyes during the withdrawal process, and add Neiguan (PC 6) when there are withdrawal symptoms such as irritability, depression, and insomnia.(**Supplementary Figure 1**)

#### Acupoint Location

Refer to the Acupuncture Point Locations in the Western Pacific Region by World Health Organization [13]. Needle Specification: 0.25 mm × 40 mm disposable sterile needles were used (purchased from Suzhou Medical Appliance Factory in China).

#### Manipulation of Acupuncture

75% medical ethanol cotton ball was used for local routine disinfection. Needles were horizontally inserted at Baihui (GV 20) and Lieque (LU 7) while vertically inserted at other acupoints with a depth of 25 to 50 mm. The mild reinforcing-reducing method was applied at all acupoints. The arrival of (needle sensation) was required and the same side Zusanli(ST 36) and Lieque(LU 7) was connected to with KWD-808 pulse electroacupuncture device (Changzhou Wujin Great Wall Medical Equipment Co., LTD.) with Continuous wave at a frequency of 15 Hz for 20 min. Meanwhile, the needles were withdrawn quickly to avoid bleeding or hematoma.

#### Frequency

Acupuncture was given 5 times a week for high-frequency acupuncture group and 3 times a week for low-frequency acupuncture group, both lasting for 8 weeks.

#### Acupuncturists

Acupuncturists in this study were all certified by Chinese Medicine Council with average acupuncture experience of 5 years; they received guidance and curriculum from the Institute of Acupuncture and Moxibustion, China Academy of Chinese Medical Sciences, to guarantee the rigor and consistency of acupuncture treatment.

### Behavioral Support

Participants were invited to join a self-help forum set up for the trial participants on WeChat, a messaging app. This was to share their experience with stopping smoking and provide mutual support via text messages. WeChat was also used for scheduling study appointments and sending appointment reminders. No other behavioral support was provided.

### Nicotine metabolic rate

In this study, 10 mL of blood was collected from the subjects’ elbow veins. The blood was centrifuged at a low speed of 3000 r/min with a centrifugal radius of 13.5 cm for 10 minutes. The plasma and blood cells were separated and stored in 10 cryopreservation tubes, which were then marked and placed in a −80°C freezer. The concentrations of cotinine and 3-hydroxycotinine in the blood samples were measured using liquid chromatography-mass spectrometry (LC-MS) by Beijing Huizhi Taikang Pharmaceutical Technology Co., Ltd. The Nicotine Metabolic Rate (NMR) was calculated as the ratio of 3-hydroxycotinine to cotinine. We chose 0.31 as the cut-off vaule based on previous studies [14, 15], which defined slow metabolizers as those with NMR ≤0.31, and normal metabolizers as those with NMR>0.31.

### Measures

At baseline, demographic and smoking history variables were collected, including age, sex, education, marital status, health status, cigarettes smoked per day, smoking duration and FTND. Objective measures included expired CO and urine cotinine test. At each follow-up, participants provided information on their smoking status, ratings of withdrawal symptoms using the Minnesota Nicotine Withdrawal Scale, use of the allocated and non-allocated study treatments. Participants were also asked whether they experienced any of the following since the previous visit: abnormal dreams, restlessness, irritability, mouth ulcers, increased appetite, dry mouth, headache, weight gain, nausea, upper respiratory tract infection, constipation, hand tremor, fatigue, insomnia, dizziness, and difficulty concentrating. CO and urine cotinine level were also collected.

### Outcomes

The primary outcome was 12-week continuous abstinence rate, assessed by urine cotinine concentrations <100 ng/mL [16]. The secondary outcome included point abstinence assessed by expiratory CO (device purchased from Bedfont Scientific Company, Maidstone, UK) of < 6 ppm [17]. Participants lost to follow-up were included as nonabstainers. Treatment adherence outcomes included attendance at monthly sessions. Other outcomes included ratings of treatments, monitoring of adverse reactions and recording of serious adverse events.

### Sample size calculation

At present, no studies have been conducted on the relationship between acupuncture for smoking cessation and nicotine metabolic rate, and there is no clear sample size to calculate reference values. In this study, we refer to the literature to calculate the difference in data of NMR on the efficacy of nicotine replacement therapy. According to the formula for calculating the sample size of the two groups of superiority test, α=0.05 and β=0.2 were taken, and 98 participants per group was calculated using PASS 15 software. Considering the 10% dropout rate, a total of 220 people was recruited.

### Statistical analysis

The measurement data are presented as means (SD). The t-test was used for comparisons which met Gaussian distribution and homogeneity of variance, whereas the nonparametric test was used for comparisons which did not meet homogeneity of variance. The categorical variables were presented with numbers (percentages), and chisquare test was used for numeral data comparisons. For primary analyses, we used the intention-to-treat approach in which participants with unknown smoking status were included as nonabstainers so that all randomized participants were included. To adjust the calculations for comparisons, OR was adjusted by age, gender, cigarettes per day (cpd), self-score of confidence in this cessation, acupuncture frequency, pre-treatment FTND score, and pre-treatment CO level. SPSS 26.0 statistical software (SPSS, Inc.) was used for statistical analysis. *P*<0.05 was regarded as statistical significance. The authors had no access to information that could identify individual participants during or after data collection.

## Results

Between September 2018 and April 2022, 258 potential participants were screened, and 220 eligible participants were randomly assigned to 1 of the 2 study arms. Altogether, 77 (70%) in the high-frequency acupuncture group and 85 (77.3%) in the low-frequency acupuncture group completed the 3-month study period. **Figure 1** showed the distribution of study dropouts over time.

**Figure 1.**
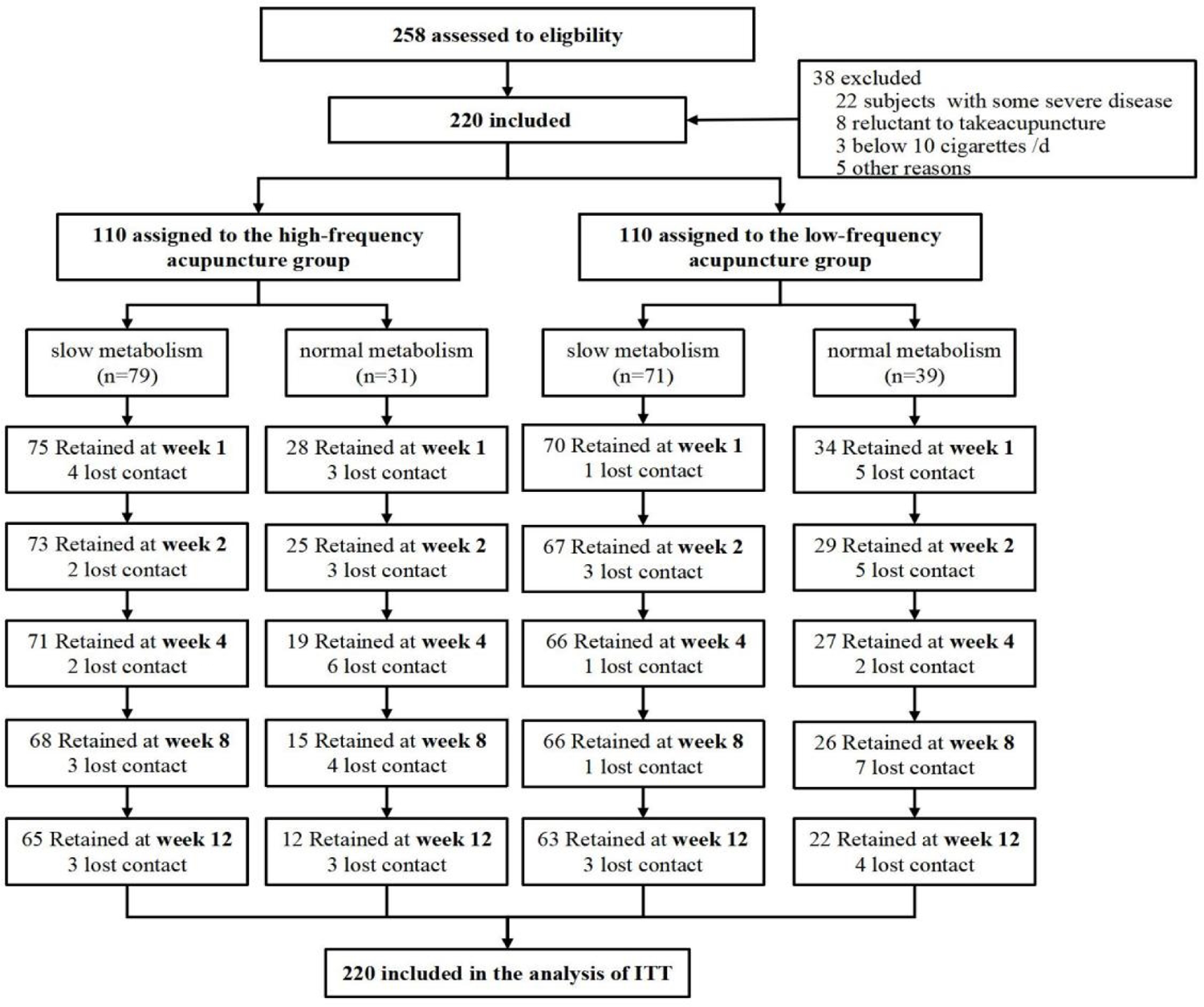
The flowchart of the study participants

Baseline characteristics of the study population were shown in **Table 1**. Overall, there were 211 male participants (95.9%), the mean (SD) age was 48.5 (11.6) years, and participants smoked on average 20.5 (10.9) cigarettes per day, with smoking duration of 27.1 (12.1) years and FTND score of 4.5 (2.3) points. No significant differences in baseline characteristics were observed between the two groups (all *P*>0.05).

**Table 1.** Baseline characteristics of study participants.

| Characteristics | High-frequency acupuncture group |  |  | Low-frequency acupuncture group |  |  | Total<br>N=220 |
| --- | --- | --- | --- | --- | --- | --- | --- |
|  | (n =110) |  |  | (n =110) |  |  |  |
|  | Slow<br>metabolizers<br>(n =79) | Normal<br>metabolizers<br>(n =31) | Total<br>(n =110) | Slow<br>metabolizers<br>(n =71) | Normal<br>metabolizers<br>(n =39) | Total<br>(n =110) |  |
| Gender |  |  |  |  |  |  |  |
| Men | 77 (97.5) | 28 (90.3) | 105 (97.5) | 69 (97.2) | 37 (94.9) | 106 (96.4) | 211 (95.9) |
| Women | 2 (2.5) | 3 (9.7) | 5 (4.5) | 2 (2.8) | 2 (5.1) | 4 (3.6) | 9 (4.1) |
| Age (year) |  |  |  |  |  |  |  |
| Less than 40 | 22 (27.8) | 10 (32.3) | 32 (29.1) | 16 (22.5) | 8 (20.5) | 24 (21.8) | 56 (25.4) |
| 41 to 50 | 22 (27.8) | 10 (32.3) | 32 (29.1) | 22 (31.0) | 15 (38.5) | 37 (33.6) | 69 (31.4) |
| 51 to 60 | 20 (25.3) | 10 (32.3) | 30 (27.3) | 20 (28.1) | 9 (23.1) | 29 (26.4) | 59 (26.8) |
| 61 and above | 15 (19.0) | 1 (3.2) | 16 (14.5) | 13 (18.3) | 7 (17.9) | 20 (18.2) | 36 (16.3) |
| Mean (SD) | 48.02±11.84 | 47.44 ±11.53 | 47.46±11.45 | 49.44±11.82 | 49.55±11.11 | 49.44±11.82 | 48.50±11.64 |
| Marriage |  |  |  |  |  |  |  |
| Single | 11 (13.9) | 6 (19.4) | 17 (15.5) | 8 (11.3) | 3 (7.7) | 11 (10.0) | 28 (12.73) |
| Married | 65 (82.3) | 24 (77.4) | 89 (80.9) | 63 (88.7) | 33 (84.6) | 96 (87.3) | 185 (84.09) |
| Separated/divorced/<br>widowed | 3 (3.8) | 1 (3.2) | 4 (3.6) | 0 (0.0) | 3 (7.7) | 3 (2.7) | 7 (3.18) |
| Education |  |  |  |  |  |  |  |
| Primary school or | 5 (6.3) | 1 (3.2) | 6 (5.5) | 3 (4.2) | 3 (7.7) | 6 (5.5) | 12 (5.45) |
| Middle and high | 41 (51.9) | 13 (41.9) | 54 (41.9) | 18(25.4) | 14 (35.9) | 32 (29.1) | 86 (39.09) |
| College and higher | 33 (41.8) | 17 (54.8) | 50 (45.5) | 50(70.4) | 22 (56.4) | 72 (65.5) | 122 (55.45) |
| Self-reported health<br>status |  |  |  |  |  |  |  |
| Poor | 9 (11.4) | 4 (12.9) | 13 (11.8) | 13(18.3) | 6 (15.4) | 19 (17.3) | 32 (14.6) |
| Average | 48 (60.8) | 13 (41.9) | 61 (55.5) | 34 (47.9) | 22 (56.4) | 56 (50.9) | 117 (53.2) |
| Good | 22 (27.8) | 14 (45.2) | 36 (32.7) | 24 (38.8) | 11 (28.2) | 35 (31.8) | 71 (32.3) |
| Cigarettes smoked<br>per day |  |  |  |  |  |  |  |
| 1-9 | 24 (30.4) | 1 (3.2) | 25 (25.5) | 21 (29.6) | 4 (10.3) | 25 (22.7) | 50 (22.7) |
| 10-19 | 41 (51.9) | 15 (48.4) | 56 (56.4) | 39 (54.9) | 21 (53.8) | 60 (54.5) | 116(52.7) |
| 20 and above | 14 (17.7) | 15 (48.4) | 29 (18.2) | 11 (15.5) | 14 (35.9) | 25 (22.7) | 54 (245) |
| Mean (SD) | 18.10±8.48 | 26.29±8.52 | 20.41±10.08 | 18.53± 9.75 | 24.26±9.75 | 20.56±11.80 | 20.49±10.95 |
| Smoking duration |  |  |  |  |  |  |  |
| 1-9 | 7 (8.9) | 4 (12.9) | 11 (10.0) | 5 (7.0) | 2 (5.1) | 7 (6.4) | 18 (8.2) |
| 10-1915 | 15 (19.0) | 5 (16.1) | 20 (18.2) | 17 (23.9) | 5 (12.8) | 22 (20.0) | 42 (19.1) |
| 20-29 | 18 (22.8) | 8 (25.7) | 26 (23.6) | 16 (22.5) | 17 (43.6) | 33 (30.0) | 59 (26.8) |
| 30 and above | 39 (49.4) | 14 (45.2) | 53 (48.2) | 33 (46.5) | 15 (38.5) | 48 (43.6) | 101 (45.9) |
| Mean (SD) | 26.57±11.36 | 21.57±11.46 | 26.47±11.35 | 27.55±12.89 | 27.60±12.37 | 27.56±12.89 | 27.05±12.09 |
| FTND |  |  |  |  |  |  |  |
| 0–3 | 27 (34.2) | 9 (29.0) | 36 (32.7) | 27 (38.0) | 18 (46.2) | 45 (40.9) | 81 (36.8) |
| 4-6 | 33 (41.8) | 15 (48.4) | 48 (43.6) | 31 (43.7) | 14 (35.9) | 45 (40.9) | 93 (42.3) |
| 7 and above | 19 (24.1) | 7 (22.6) | 26 (23.6) | 13 (18.3) | 7 (17.9) | 20 (18.2) | 46 (20.9) |
| Mean (SD) | 4.77±2.30 | 4.79±2.30 | 4.41±2.46 | 4.20±2.32 | 3.23±2.34 | 4.55±2.19 | 4.48±2.32 |

With 0.31 as the NMR cut-off value, 70 (31.8%) were divided as normal metabolizers and 150 (68.2%) as slow metabolizers. The average of 18.31 (9.91) cigarettes per day for the slow metabolizers was significantly lower than that of 25.16 (11.65) for the normal metabolizers (*P*<0.05). No other significant differences in baseline characteristics were observed (*P>0.05*) (**Supplementary Table 1**).

The validated 3-month sustained abstinence rates were 17.3% (19/110) in the high-frequency acupuncture group and 8.2% (9/110) in the low-frequency acupuncture group, with high-frequency acupuncture superior to low-frequency acupuncture (OR=3.89, 95% CI: 1.36, 8.11). In addition, the validated 2-month abstinence rates were 29.1% (32/110) in the high-frequency acupuncture group and 26.4% (29/110) in the low-frequency acupuncture group, with high-frequency acupuncture non-inferior to low-frequency acupuncture (OR=1.51, 95% CI: 0.72, 3.12) (**Table 2**).

**Table 2.** Validated abstinence rates by acupuncture frequency at different time points.

| Primary outcome | High-frequency<br>acupuncture group (n<br>=110) | Low-frequency<br>acupuncture group (n<br>=110) | OR (95 CI) |
| --- | --- | --- | --- |
| 2 months (End of Treatment) | 29.1 (32/110) | 26.4 (29/110) | 1.51 (0.72, 3.12) |
| 3 months (End of Study) | 17.3 (19/110) | 8.2 (9/110) | 3.89 (1.36, 8.11) |

**Note:** the primary outcome was self-reported continuous abstinence rate verified by urine cotinine concentrations <100 ng/mL; OR was adjusted by age, gender, self-reported health status, cigarettes per day (cpd), smoking duration, self-score of confidence in this cessation, acupuncture frequency, pre-treatment FTND score.

In slow metabolizers, the validated 3-month sustained abstinence rate was 19.0% (15/79) in the high-frequency acupuncture, which was significantly higher than 7.0% (5/71) in low-frequency acupuncture (OR=3.53, 95% CI: 1.03, 6.13). In normal metabolizers, the validated 3-month sustained abstinence rates were 12.9% (4/31) in the high-frequency acupuncture group and 10.3% (4/39) in the low-frequency acupuncture group, with high-frequency acupuncture non-inferior to low-frequency acupuncture (OR=1.11, 95% CI: 0.24, 5.17)). (**Table 3**)

**Table 3.** Validated abstinence rates by NMR at different time points.

| Outcome | Slow metabolizers (n =150) |  |  | Normal metabolizers (n =70) |  |  |
| --- | --- | --- | --- | --- | --- | --- |
|  | High-frequency acupuncture group(n=71) | Low-frequency acupuncture group(n=110) | OR (95% CI) | High-frequency acupuncture group (n=110) | Low-frequency acupuncture group(n=110) | OR (95% CI) |
| 2 months | 29.1 (23/79) | 26.8 (19/71) | 1.43 (0.57, 3.63) | 29.0 (9/31) | 25.6 (10/39) | 1.06 (0.36, 3.16) |
| 3 months | 19.0 (15/79) | 7.0 (5/71) | 3.53 (1.03, 6.13) | 12.9 (4/31) | 10.3 (4/39) | 1.11 (0.24, 5.17) |

**Note:** the primary outcome was self-reported continuous abstinence rate verified by urine cotinine concentrations <100 ng/mL; OR was adjusted by age, gender, cigarettes per day (cpd), self-score of confidence in this cessation, acupuncture frequency, pre-treatment FTND score, and pre-treatment CO level.

Treatment adherence was similar in the two study arms (**Table 4**). Nearly all the participants set target quit date (95.9%), and all used at least 1 acupuncture treatment. Besides, no use of any of allocated treatment was recorded in the two study arms after the initial 3 months.

**Table 4.** Treatment adherence, % (N)

| Treatment adherence | High-frequency acupuncture group (n =110) | Low-frequency acupuncture group (n =110) | P | Total (n=220) |
| --- | --- | --- | --- | --- |
| Set target quit date (TQD) | 96.4 (106/110) | 95.5 (105/110) | 0.733 | 95.9 (211) |
| Used at least 1 acupuncture treatment | 100.0 (110/110) | 100.0 (110/110) | — | 100.0 (220/220) |
| Used at least 80% of acupuncture treatment | 82.7 (91) | 86.4 (95) | 0.456 | 84.5 (186) |
| Using allocated treatment during follow-up | 0.0 (0) | 0.0 (0) | — | 0.0 (0) |
| Used non-allocated treatment at any time point | 6.4 (7) | 5.5 (6) | 0.7749 | 5.9 (133) |

Adverse reactions were infrequent (5 participants in the high-frequency group and 3 participants in the low-frequency group), which included primarily local hematoma. No serious adverse events were reported in any of the two study arms.

## Discussion

To the best of our knowledge, this was the first randomized controlled trial to use NMR as an informed biomarker to improve the efficacy of acupuncture for smoking cessation, offering significant insights for future research and clinical practice. Our findings demonstrated that high-frequency acupuncture was more effective for slow metabolizers, while low-frequency acupuncture efficacy did not differ significantly between slow and normal metabolizers. This suggests that NMR might be a crucial biomarker for tailoring acupuncture treatments, particularly for slow metabolizers.

The overall abstinence rates in our study align with previous researches [18–20] indicating that acupuncture is an effective intervention for smoking cessation. Our findings support acupuncture’s role as an effective alternative to conventional smoking cessation therapies such as nicotine replacement therapy (NRT) and varenicline. However, the abstinence rates observed in our study were relatively modest compared to some pharmacological interventions [21], suggesting that acupuncture might be best utilized in conjunction with other treatments for optimal results.

The role of NMR as a biomarker in our study provided novel insights into the personalization of smoking cessation strategies. Slow metabolizers (NMR≤0.31) exhibited higher abstinence rates with high-frequency acupuncture compared to low-frequency acupuncture. This finding suggests that slow metabolizers might benefit more from intensive acupuncture interventions. Conversely, no significant differences were observed between high-frequency and low-frequency acupuncture among normal metabolizers (NMR>0.31), indicating that other factors might influence their cessation outcomes. These results[22,23] also align with previous studies showing that normal metabolizers have lower success rates with standard cessation therapies due to their rapid nicotine clearance.

Furthermore, the interaction between NMR and acupuncture points to a complex biochemical and physiological process. Nicotine metabolism, primarily governed by the CYP2A6 enzyme, affects how quickly nicotine is processed in the body [24]. Slow metabolizers, with lower CYP2A6 activity, maintain higher nicotine levels for extended periods, potentially experiencing less severe withdrawal symptoms. High-frequency acupuncture might further alleviate withdrawal symptoms and cravings in these individuals by modulating neurotransmitter release and enhancing endorphin levels [25], thereby supporting smoking cessation.

In contrast, normal metabolizers break down nicotine rapidly, leading to more pronounced withdrawal symptoms and cravings, which might not be adequately addressed by acupuncture alone. This highlights the need for additional support, possibly integrating pharmacological treatments like nicotine replacement therapy or varenicline, to improve cessation outcomes in this group.

The identification of NMR as a predictive biomarker for acupuncture efficacy could revolutionize personalized smoking cessation strategies. By tailoring acupuncture frequency and intensity based on an individual’s metabolic rate, healthcare providers can enhance treatment efficacy, reduce relapse rates, and optimize resource allocation. This approach aligns with the broader trend towards personalized medicine, emphasizing the customization of healthcare interventions based on individual characteristics.

The high adherence rates and low incidence of adverse reactions further underscore acupuncture’s feasibility and safety as a smoking cessation tool. Participants’ engagement with the WeChat support group also highlights the potential of integrating digital health interventions to enhance treatment adherence and support[26].

Despite the promising results, our study has several limitations. First, the sample size, while calculated based on existing literature, might not fully capture the variability in NMR and its impact on acupuncture efficacy. Future studies with larger sample sizes are needed to validate our findings. Second, the study population consisted predominantly of male participants, which limits the generalizability of our results to female smokers. Third, the study relied on self-reported smoking status verified by urine cotinine concentrations which, despite being standard practice, may introduce some biases[27]. Fourth, another external event affecting the trial was the COVID-19 pandemic. The lockdowns made the collection of CO readings and urine cotinine difficult. This reduced the validated quit rates, but both study arms were affected equally. This event was the likely reason for quit rates being lower than expected in our power calculations. Another limitation is the potential for unmeasured confounding variables that could influence both NMR and smoking cessation outcomes. While we adjusted for known confounders, there may be other factors not accounted for that could impact the relationship between NMR and acupuncture efficacy. Future research should aim to identify and control for these variables to provide a clearer understanding of the mechanisms at play.

In conclusion, our study provides pioneering evidence that NMR can be used to optimize acupuncture treatments for smoking cessation, particularly among slow metabolizers. Further research should explore the underlying mechanisms of NMR’s influence on acupuncture efficacy and extend these findings to more diverse populations. Integrating acupuncture with other behavioral and pharmacological interventions may also enhance smoking cessation outcomes, offering a comprehensive approach to tackling this significant public health issue.

### Primary funding

China Capital Health Development Special Project (2024-2-4372); China Youth Qihuang Fund Special Project (11055); Science and Technology Innovation Project of China Academy of Chinese Medical Sciences (CI2021A03506); China Capital Health Development Special Project (2023-1-4281)

## Data Availability

Researchers who meet the criteria for access to confidential data can access the data from the corresponding author.

## Contributors

All authors were involved in the planning of the study, literature review, data collection, interpretation of the findings, and manuscript preparation. Wang YY, Yang JS and Liu Z conceived and designed the study. Wang YY supervised the study. Chen SM, Ji J, Liu ZY, Yang L,Chai X,Wu Y, Chang J,Tan CR,and Zhang M contributed to the acquisition of data. Wang YY, Chen SM and Liu Z drafted the report. Chen SM did the statistical analysis. All authors revised the report and approved the final version before submission. Wang YY is the guarantor and attest that all listed authors meet authorship criteria and that no others meeting the criteria have been omitted.

## Data sharing statement

Data will be available upon reasonable request to the corresponding author immediately following publication to anyone wishing to access the data.

## Acknowledgments

The authors would like to thank the study participants, and appreciate every supporter who contributed to this study.

---

**Supplementary Figure 1.**
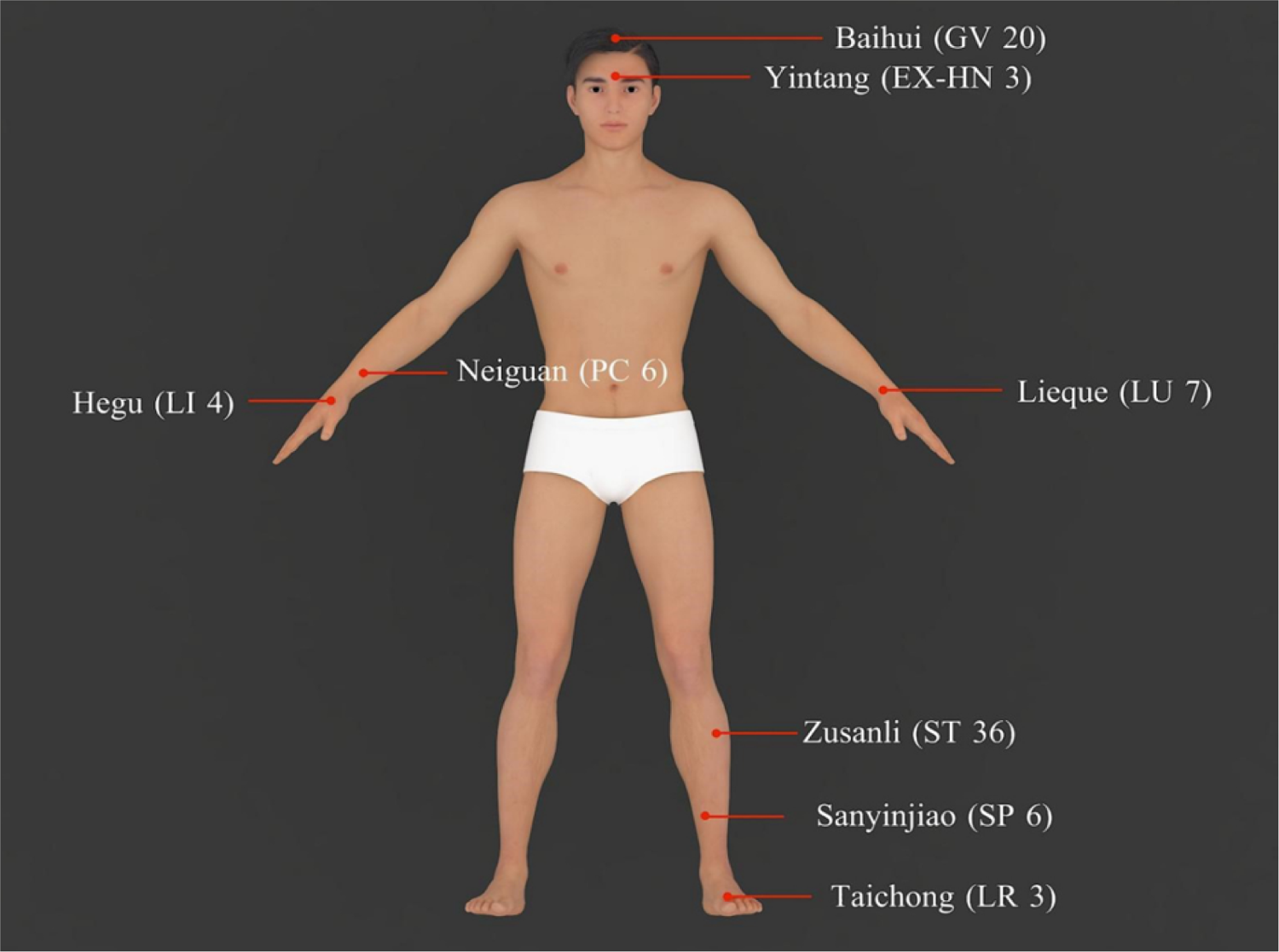
The acupoints used in the study The main acupoints: Baihui (GV 20), Lieque (LU 7), Hegu (LI 4), Zusanli (ST 36), Sanyinjiao (SP 6), Taichong (LR 3) with Yintagb(EX-HN 3), Neiguan(PC 6)

**Supplementary Table 1.**
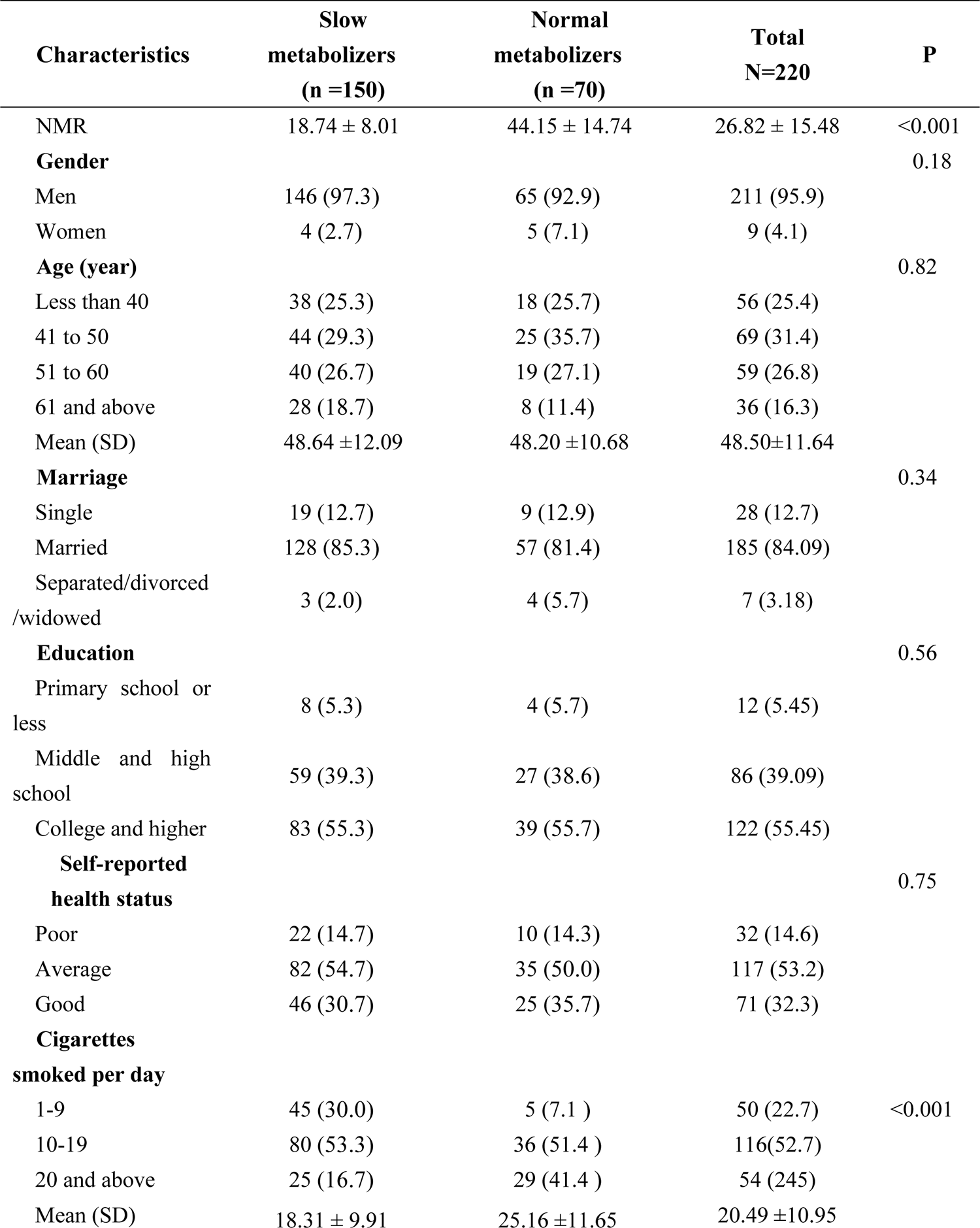
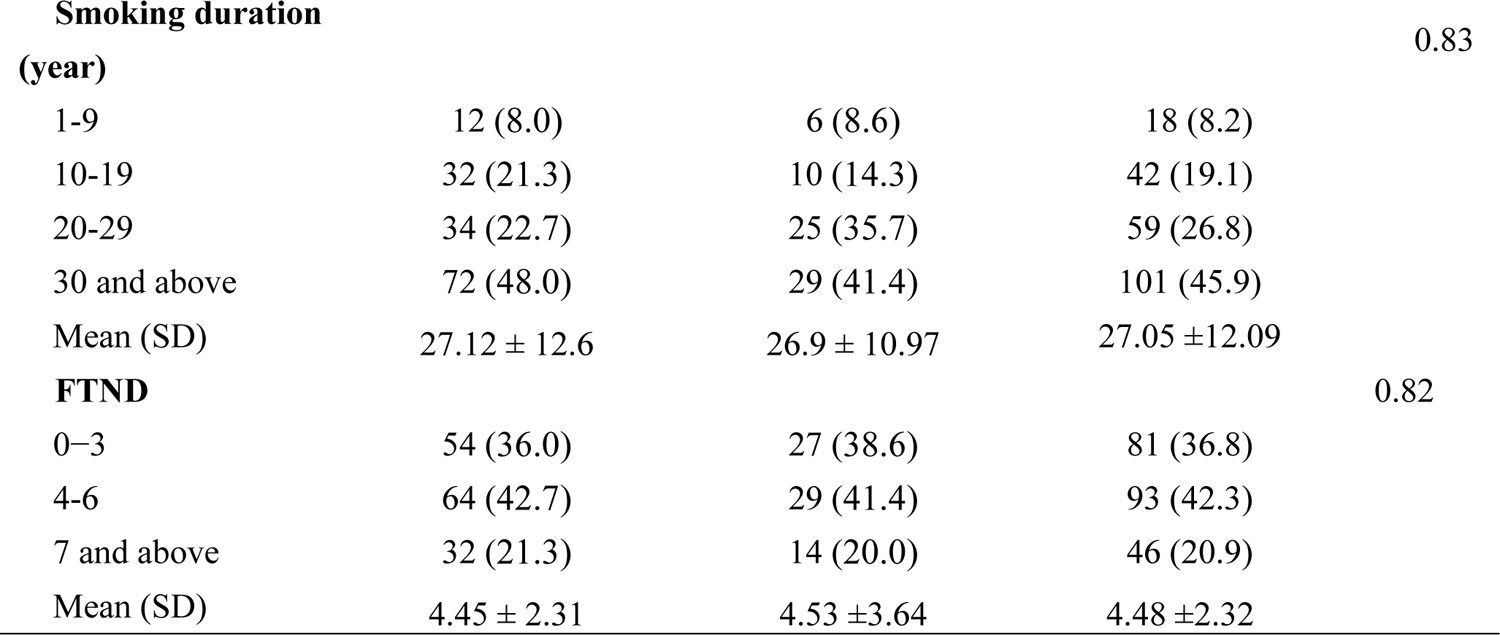
Baseline characteristics of study participants by NMR.

